# Exploring the Association Between Urinary Incontinence and Depression Based on a Series of Large-Scale National Health Studies in Turkiye

**DOI:** 10.1101/2025.01.10.25320344

**Authors:** Muhammed Furkan Dasdelen, Zehra Betul Dasdelen, Furkan Almas, Beyza Cokkececi, Pilar Laguna, Jean de la Rosette, Mehmet Kocak

## Abstract

Urinary incontinence (UI) and depression are prevalent conditions affecting millions globally and are significantly associated with various demographic, health, and socio-economic factors. This study examines the trends and associations of UI and depression over a 14-year period using nationwide data. We analyzed data from the Turkish Health Studies Surveys conducted in seven years between 2008 and 2022, including 125,279 participants aged 15 and older, excluding those with incomplete key health data. Variables included chronic conditions, BMI, depression severity (assessed by PHQ-8), socio-economic status, and lifestyle factors. Univariable and multivariable logistic regression models were used to investigate associations between UI and various risk factors over time. The prevalence of UI and depression fluctuated over the 14-years, with a significant increase observed in 2014. Multivariate analysis confirmed a strong and consistent association between UI and depression across genders and age groups, even after adjusting for confounders. Higher depression severity increased the odds of experiencing UI. Age, multiple comorbidities, higher BMI, and lower socio-economic status were associated with an increased likelihood of UI. Obesity was a significant risk factor for UI in females but not in males. Urban living and higher education levels were inversely associated with UI. The simultaneous rise in UI and depression in 2014 may be linked to socioeconomic changes during that period. The findings suggest a robust link between UI and depression, influenced by a complex interplay of health, demographic, and socio-economic factors, needing prospective studies to further investigate the causal pathway of these associations.

## Introduction

Urinary incontinence (UI) is a common and often distressing condition that affects over 400 million people worldwide (1). While it is often assumed to be a concern primarily for women and the elderly, UI can be seen across all age groups and sexes (2). Studies indicate that up to 25-45% of women experience some degree of urinary incontinence, with the incidence rising with age (2; 3). In men, the prevalence is lower, estimated at around 2-11%, but also increases with age (2; 4; 5). Cross-sectional and longitudinal studies have found associations between UI and certain risk factors, such as increased BMI (6; 7; 8), childbirth (9; 8), menopause (10; 8), and chronic conditions including diabetes, cardiorespiratory disorders, musculoskeletal problems (11; 12; 13). These increasing risk factors with age elevate the likelihood of developing UI in the elderly (11; 8). Regardless of age, UI remains bothersome for all, leading to a lack of self-confidence, embarrassment, disturbances in work or social life, and adverse psychosocial outcomes, including depression, social isolation, and anxiety (14; 15). Previous cross-sectional studies showing a strong association between UI, depression, and anxiety have been reported in European and Western countries (16; 17; 18; 19; 20; 21). Although the majority of these studies have been conducted on women, the association has also been demonstrated in men (16; 18). The causal direction of this relationship was better observed through longitudinal studies. Prospective studies found that both depression and anxiety are risk factors for UI (22; 23; 24; 25; 26; 27; 28; 29), and UI can be a predictor of both (22; 23; 15; 28), suggesting a potential bidirectional causality. Further research is needed to better understand this relationship, considering possible confounders such as demographics, environmental factors, and comorbidities in different populations. In our previous study, we examined the relationship between depression, anxiety, and urinary incontinence in the presence of various comorbidities (30). We found that the prevalence of urinary incontinence increases as the level of psychological discomfort rises amongst the Turkish population. This relationship might be a direct or indirect cause-and-effect relationship, which can be better observed through longitudinal studies. In the present study, we examined data from the Turkiye Health Interview Survey, collected over seven different years between 2008 and 2022. It includes a total of 125,279 participants representing the whole population at the time of collection. Our first aim is to discover the trend of urinary incontinence and depression prevalence over this 14-year period and explore external factors influencing this relationship. Our second aim is to investigate the consistency of the association between depression and urinary incontinence in each cross-sectional dataset, considering the comorbidities and characteristics of participants.

## Methods

### Data Collection

As adopted modules from the Eurostat European Health Interview Survey (EHIS) and conducted as a part of EHIS waves, Turkiye Health Survey was carried out first in 2008 and was implemented every two years until 2016, and every three years afterwards in 2019 and 2022. The data collection technique has been detailed in earlier literature (30; 31; 32). In summary, a stratified two-stage cluster sampling approach was employed. For external stratification, differentiation between rural and urban areas was utilized. The initial stage involved the random selection of blocks proportional to the size of clusters. In the second stage, household addresses were selected systematically and randomly from each chosen cluster. The sampling spanned all geographical regions and included every resident in Turkiye, except those in institutional settings (such as military personnel, dormitory inhabitants, long-term hospital patients, elderly home residents, etc.) and very small settlements unable to provide a sufficient number of sample households (like small villages or hamlets). The total sample size has varied, from 7,910 in 2008 to 11,179 households in 2022. Interviews were conducted face-to-face, with the first survey taking place in April 2008. Subsequent surveys occurred over one month in May-June during 2010 and 2012, and over three months in August-October during 2014 and 2016, and September-December in 2019 and 2022. The participants provided informed consent before joining the study. Ethical approval was granted by the Istanbul Medipol University Ethics Committee (Application number: 10840098-604.01.01-E.53819). All procedures were carried out in compliance with the Declaration of Helsinki. The survey’s objective is to collect data on the health indicators of Turkiye periodically, enabling the monitoring of changes in the population’s health status and its determinants. According to the Eurostat methodology, although some questions were modified over the years, the survey consistently targeted individuals aged 0-14 and 15+. For those over 15, the survey included three primary modules: health status, health care services, and health determinants. The health status module encompasses diseases and chronic conditions, accidents, injuries, physical and sensory impairments, personal care activities, pain, and mental health issues. The health care services module covers the usage of inpatient and outpatient services, medications, and preventive measures. Lastly, the health determinants module includes measurements of weight, height, physical activity, dietary habits, and the use of alcohol and tobacco.

### Variables and Categorization

Participants self-reported their chronic conditions, responding with ‘Yes’, ‘No’, ‘Do not know’, or ‘Refuse to answer’. The options ‘Do not know’ and ‘Refuse to answer’ were omitted after 2012. Participants were also given the option not to answer a specific question. Those who answered ‘Yes’ were considered to have the disease. Individuals with any of the following cardiovascular conditions: myocardial infarction, hypertension, chronic heart failure, or coronary artery disease, were categorized as having a cardiac disease. The PHQ-8 scale was utilized to assess the severity of depression among those who reported depression. The distinction between urban and rural lifestyles was only included in the years 2008, 2010, and 2012. Regional categorization was based on the Nomenclature of Territorial Units for Statistics (NUTS)-2, which divides major socio-economic regions. Education levels were classified according to the International Standard Classification of Education (ISCED) 2011 and Eurostat’s guidelines. ISCED 2011 levels 0-2 were classified as low education, levels 3-4 as medium education, and levels 5-8 as high education. For body mass index (BMI), categories were defined as follows: underweight for a BMI less than 18.5, normal weight for a BMI ranging from 18.5 to 24.9, overweight for a BMI from 25.0 to 29.9, and obese for a BMI of 30.0 and above.

### Eligibility Criteria

Initially, individuals under 15 years of age were excluded (Figure 1). Additionally, we excluded cases with missing values for diseases such as asthma, COPD, myocardial infarction, hypertension, coronary artery disease, stroke, osteoarthritis, diabetes, cirrhosis, urinary incontinence, or depression.

**Figure 1.**
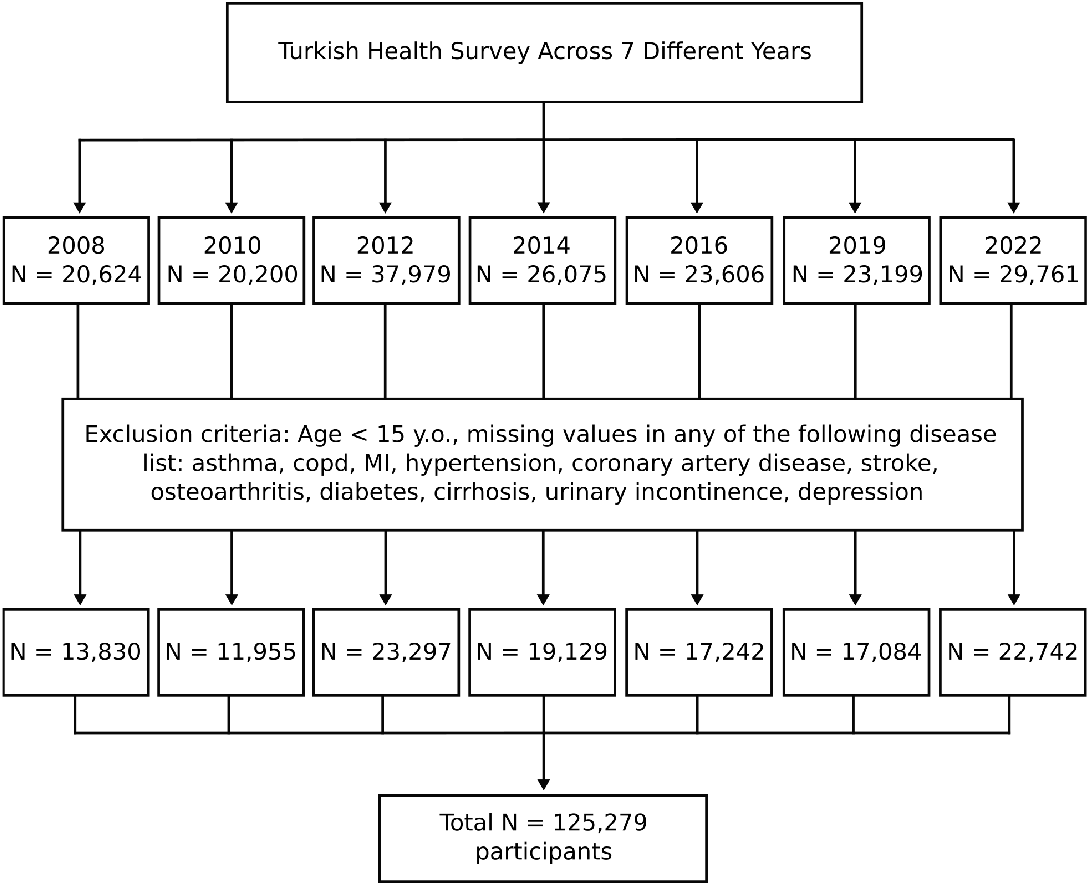
Flow chart of the study. A total of 125,279 participants were included in the study, after applying the exclusion criteria to the participants from 7 different years. For each year, separate multivariable logistic regression models were utilized to evaluate association of urinary incontinence and independent variables.

## Statistical Analysis

We utilized univariable and multivariable logistic regression models to examine the relationship between urinary incontinence (UI) and various factors. For biological and demographic factors, we employed separate multivariable logistic regression models for each year, with UI as a binary dependent variable and other chronic conditions, age, and BMI as independent variables. For lifestyle-socioeconomic factors and depression severity, we initially applied univariable logistic regression and subsequently adjusted for age and chronic conditions using multivariable logistic regression. We used the Chi-squared test to analyze prevalence differences according to income levels. For UI and depression prevalence trends over years, we fit two different interrupted time series regression models. We assumed an event interrupting the natural trend of the UI or depression prevalence between 2012 and 2014. We wrote the prevalence as a function of time:

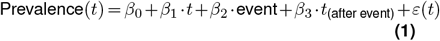

In Equation Eq. (1):

- *β*_0_ is the baseline level of the prevalence.
- *β*_1_ represents the trend of prevalence before the event.
- *β*_2_ captures the immediate level change in prevalence right after the event.
- *β*_3_ reflects the change in the trend of prevalence after the event.

The variable event is a binary indicator, where it equals 0 for times *t* before the event and 1 for times *t* after the event. The term *ε*(*t*) denotes the error term or residuals at time *t*. Prais-Winsten estimation from prais package in R was utilized to take care of autocorrelation. For a detailed analysis of the BMI and UI relationship, we conducted restricted cubic spline (RCS) analysis separately for genders to explore potential non-linear associations. The data from all years were concatenated and people with missing BMI values were eliminated (Number of eliminated people: n = 1226 for males, n = 3833 for females). The analysis conducted on 55,135 males and 65,085 females. Knots were placed at the 5th, 25th, 50th, 75th, and 95th percentiles of the BMI distribution within each gender subgroup to capture the variability in BMI adequately. We included the transformed BMI variables in a multivariable logistic regression model with UI as the dependent variable and adjusted for age and chronic conditions In generating predictions from the model, we set age to its mean value and the binary comorbidity variables to 1. We experimented with setting the comorbidity variables to 0, the median value, the mean value or 1. All settings produced similar graph shapes but varied in the odds values of UI and the confidence intervals. In this study, we preferred setting the comorbidity variables to 1 as it provided clearer visualizations. A p-value of less than 0.05 was considered statistically significant throughout the study. Bonferroni correction was applied for multiple comparisons and indicated where applicable. Statistical analysis and visualizations were conducted using Python v3.10. For pooling odds ratios from different years, we used random-effect models provided by the metafor package in R. The key programming codes for this analysis have been made available online as Supplementary Data.

## Results

After applying the exclusion criteria across data from seven different years, a total of 125,279 participants were included in the study. The number of participants per year was as follows: 13,830 in 2010, 11,955 in 2011, 23,297 in 2012, 19,129 in 2013, 17,242 in 2014, 17,084 in 2015, and 22,742 in 2016. The highest number of participants was recorded in 2012, while the lowest was in 2010 (Figure 1). The demographic and medical characteristics of the participants in each year are demonstrated in Table 1. Notably, gender distribution showed a consistent pattern, with females comprising over half of the participants each year, peaking at 58.36% in 2010. Moreover, age distribution revealed a significant decrease in the youngest age group (15-24 years) over time, from 19.93% in 2008 to 16.82% in 2022, while the proportion of participants with age more than 55 was increased from 20.98% in 2008 to 27.76% in 2022. The prevalence of urinary incontinence (UI) showed significant variation, peaking at 8.80% in 2019, up from 4.48% in 2008, before slightly decreasing to 5.58% in 2022. A dramatic rise was also observed between 2012 and 2014. Concurrently, depression rates also varied markedly, with a notable peak in 2014 (11.72%) compared to other years, and a decrease to 7.20% in 2022. Furthermore, other health conditions, such as asthma, COPD, MI, coronary heart disease, hypertension, cardiac diseases, osteoarthritis, diabetes mellitus, and cirrhosis also displayed a similar rise in 2014. On the other hand, stroke presented a notable decrease in the same year. In the following years, asthma, MI, and osteoarthritis had increasing trends till 2019 while, diabetes mellitus had a stable rise up to date. These increases may be attributed to an aging population and lifestyle changes. However, the prevalence of COPD, coronary heart disease, cardiac diseases, and cirrhosis consistently descended after 2014. In terms of education, the proportion of individuals in the low ISCED category consistently decreased, along with an increase in the medium and high categories, indicating an overall upward trend in the education level of the population. The marital status distribution demonstrated a quite steady track. On the contrary, the rural lifestyle had lost its popularity over the years, suggesting the ongoing rural depopulation of Turkiye (33). The prevalence of UI and depression was further analyzed in different subgroups with varying comorbidities (Figure 2). It is highlighted that both UI and depression were increased in parallel with the age and the number of comorbidities. Participants aged 50 years and above with multiple comorbidities, including diabetes, cardiac disease, stroke, and osteoarthritis, exhibited the highest prevalence rates of both UI and depression, promoting the statement above. Also, this uptrend is particularly pronounced in participants with a combination of diabetes, cardiac disease, stroke, and osteoarthritis. Remarkably, gender-specific data revealed that females consistently report higher rates of UI and depression compared to males across the majority of years and subgroups. We next analyzed the associations of various factors with UI using logistic regression models. First, we evaluated age, BMI and chronic conditions which are all part of biological components. After applying separate multivariable logistic regression in each year, we pooled the odds ratios for each independent variable as represented in Figure 3. Detailed odds ratios for each year were included in Supplementary Figure 1. Multivariate logistic regression analysis revealed a strong and consistent relationship between UI and depression across multiple years for both genders (Figure, Supplementary Figure 1). The pooled odds ratio of depression was 2.86 [2.53-3.24, 95%CI] in females and 2.80 [2.10-3.72 95%CI] in males, indicating a significant association. Furthermore, people with higher depression severity had higher odds of UI (Table 2). Aging remained a primary contributor to UI, with pooled odds ratios of 1.54 [1.43-1.65, 95%CI] for females and 2.15 [1.95-2.36 95%CI] for males. Additionally, several other chronic conditions including cardiac diseases, stroke, diabetes mellitus, osteoarthritis, COPD, asthma, and cirrhosis revealed significant associations with UI in both genders. Interestingly, males who were overweight compared to those of normal weight had lower odds of having UI.

**Table 1.** Characteristics of participants in each year.

| Variable | 2008<br>(N=13830) | 2010<br>(N=11955) | 2012<br>(N=23297) | 2014<br>(N=19129) | 2016<br>(N=17242) | 2019<br>(N=17084) | 2022<br>(N=22742) |
| --- | --- | --- | --- | --- | --- | --- | --- |
| **Gender** |  |  |  |  |  |  |  |
| • Female | 7475<br>(54.05%) | 6977<br>(58.36%) | 13413<br>(57.57%) | 10408<br>(54.41%) | 9574<br>(55.53%) | 9300<br>(54.44%) | 11771<br>(51.76%) |
| • Male | 6355<br>(45.95%) | 4978<br>(41.64%) | 9884<br>(42.43%) | 8721<br>(45.59%) | 7668<br>(44.47%) | 7784<br>(45.56%) | 10971<br>(48.24%) |
| **Age** |  |  |  |  |  |  |  |
| • 15-24 | 2757<br>(19.93%) | 2378<br>(19.89%) | 4626<br>(19.86%) | 3388<br>(17.71%) | 2905<br>(16.85%) | 2730<br>(15.98%) | 3825<br>(16.82%) |
| • 25-34 | 3158<br>(22.83%) | 2424<br>(20.28%) | 4541<br>(19.49%) | 3661<br>(19.14%) | 3006<br>(17.43%) | 3070<br>(17.97%) | 4077<br>(17.93%) |
| • 35-44 | 2741<br>(19.82%) | 2298<br>(19.22%) | 4447<br>(19.09%) | 3768<br>(19.70%) | 3444<br>(19.97%) | 3395<br>(19.87%) | 4628<br>(20.35%) |
| • 45-54 | 2271<br>(16.42%) | 2080<br>(17.40%) | 3983<br>(17.10%) | 3332<br>(17.42%) | 3007<br>(17.44%) | 2918<br>(17.08%) | 3901<br>(17.15%) |
| • 55-64 | 1501<br>(10.85%) | 1411<br>(11.80%) | 2870<br>(12.32%) | 2555<br>(13.36%) | 2368<br>(13.73%) | 2513<br>(14.71%) | 3167<br>(13.93%) |
| • 65-74 | 876<br>(6.33%) | 855<br>(7.15%) | 1778<br>(7.63%) | 1498<br>(7.83%) | 1545<br>(8.96%) | 1590<br>(9.31%) | 2183<br>(9.60%) |
| • 75+ | 526<br>(3.80%) | 509<br>(4.26%) | 1052<br>(4.52%) | 927<br>(4.85%) | 967<br>(5.61%) | 868<br>(5.08%) | 961<br>(4.23%) |
| **Chronic conditions** |  |  |  |  |  |  |  |
| • Asthma | 692<br>(5.00%) | 659<br>(5.51%) | 1270<br>(5.45%) | 1628<br>(8.51%) | 1499<br>(8.69%) | 1665<br>(9.75%) | 1872<br>(8.23%) |
| • COPD | 299<br>(2.16%) | 525<br>(4.39%) | 702<br>(3.01%) | 1616<br>(8.45%) | 1369<br>(7.94%) | 1373<br>(8.04%) | 1508<br>(6.63%) |
| • Myocardial infarction | 260<br>(1.88%) | 150<br>(1.25%) | 240<br>(1.03%) | 421<br>(2.20%) | 422<br>(2.45%) | 434<br>(2.54%) | 561<br>(2.47%) |
| • Coronary disease | 889<br>(6.43%) | 599<br>(5.01%) | 1113<br>(4.78%) | 1729<br>(9.04%) | 1294<br>(7.50%) | 1338<br>(7.83%) | 1494<br>(6.57%) |
| • Hypertension | 2196<br>(15.88%) | 1843<br>(15.42%) | 3647<br>(15.65%) | 3535<br>(18.48%) | 3269<br>(18.96%) | 3167<br>(18.54%) | 3831<br>(16.85%) |
| • Any cardiac disease | 2753<br>(19.91%) | 2243<br>(18.76%) | 4326<br>(18.57%) | 4407<br>(23.04%) | 3865<br>(22.42%) | 3867<br>(22.64%) | 4589<br>(20.18%) |
| • Stroke | 165<br>(1.19%) | 155<br>(1.30%) | 248<br>(1.06%) | 165<br>(0.86%) | 167<br>(0.97%) | 141<br>(0.83%) | 219<br>(0.96%) |
| • Osteoarthritis | 1782<br>(12.89%) | 1160<br>(9.70%) | 1707<br>(7.33%) | 1773<br>(9.27%) | 1608<br>(9.33%) | 2147<br>(12.57%) | 1889<br>(8.31%) |
| • Diabetes | 897<br>(6.49%) | 857<br>(7.17%) | 1836<br>(7.88%) | 1996<br>(10.43%) | 1878<br>(10.89%) | 1961<br>(11.48%) | 2743<br>(12.06%) |
| • Cirrhosis | 150<br>(1.08%) | 64 (0.54%) | 99 (0.42%) | 340<br>(1.78%) | 292<br>(1.69%) | 294<br>(1.72%) | 301<br>(1.32%) |

**Table 2.** Lifestyle and social determinants of urinary incontinence.

| Independent variables | Odds ratio [95%CI] | P value* | Adjusted odds ratio**<br>[95%CI] | P value* |
| --- | --- | --- | --- | --- |
| **Female** |  |  |  |  |
| **Lifestyle** |  |  |  |  |
| • Rural | Reference | Reference | Reference | Reference |
| • Urban | 0.64 [0.53-0.77] | <0.01 | 0.77 [0.63-0.93] | 0.01 |
| **Education** |  |  |  |  |
| • Low | Reference | Reference | Reference | Reference |
| • Medium | 0.22 [0.19-0.25] | <0.01 | 0.57 [0.50-0.66] | <0.01 |
| • High | 0.25 [0.21-0.30] | <0.01 | 0.43 [0.37-0.52] | <0.01 |
| **Marital status** |  |  |  |  |
| • Single/divorced/widowed | Reference | Reference | Reference | Reference |
| • Married | 0.92 [0.83-1.03] | 0.14 | 1.01 [0.90-1.13] | 0.75 |
| **Depression Severity (PHQ-8)** |  |  |  |  |
| • None | Reference | Reference | Reference | Reference |
| • Mild | 2.69 [2.38-3.04] | <0.01 | 2.14 [1.86-2.47] | <0.01 |
| • Moderate | 4.31 [3.72-4.99] | <0.01 | 3.47 [2.89-4.18] | <0.01 |
| • Moderately severe | 7.13 [6.10-8.34] | <0.01 | 4.97 [4.03-6.13] | <0.01 |
| • Severe | 9.14 [7.35-11.38] | <0.01 | 3.83 [3.03-4.84] | <0.01 |
| **Male** |  |  |  |  |
| **Lifestyle** |  |  |  |  |
| • Rural | Reference | Reference | Reference | Reference |
| • Urban | 0.43 [0.34-0.54] | <0.01 | 0.63 [0.49-0.80] | <0.01 |
| **Education** |  |  |  |  |
| • Low | Reference | Reference | Reference | Reference |
| • Medium | 0.18 [0.15-0.21] | <0.01 | 0.55 [0.47-0.65] | <0.01 |
| • High | 0.27 [0.22-0.32] | <0.01 | 0.54 [0.45-0.65] | <0.01 |
| **Marital status** |  |  |  |  |
| • Single/divorced/widowed | Reference | Reference | Reference | Reference |
| • Married | 2.21 [1.91-2.57] | <0.01 | 0.71 [0.61-0.83] | <0.01 |
| **Depression Severity (PHQ-8)** |  |  |  |  |
| • None | Reference | Reference | Reference | Reference |
| • Mild | 2.78 [2.40-3.22] | <0.01 | 2.34 [1.96-2.79] | <0.01 |
| • Moderate | 4.03 [3.36-4.83] | <0.01 | 3.69 [2.90-4.70] | <0.01 |
| • Moderately severe | 6.49 [5.35-7.86] | <0.01 | 4.45 [3.35-5.90] | <0.01 |
| • Severe | 14.23 [10.92-18.53] | <0.01 | 6.91 [5.12-9.32] | <0.01 |

\* After Bonferroni correction, P value less than 0.125 was considered significant. \*\*Adjusted to age and following comorbidities: depression,
asthma, COPD, cardiac diseases, stroke, osteoarthritis, diabetes, cirrhosis.

**Figure 2.**
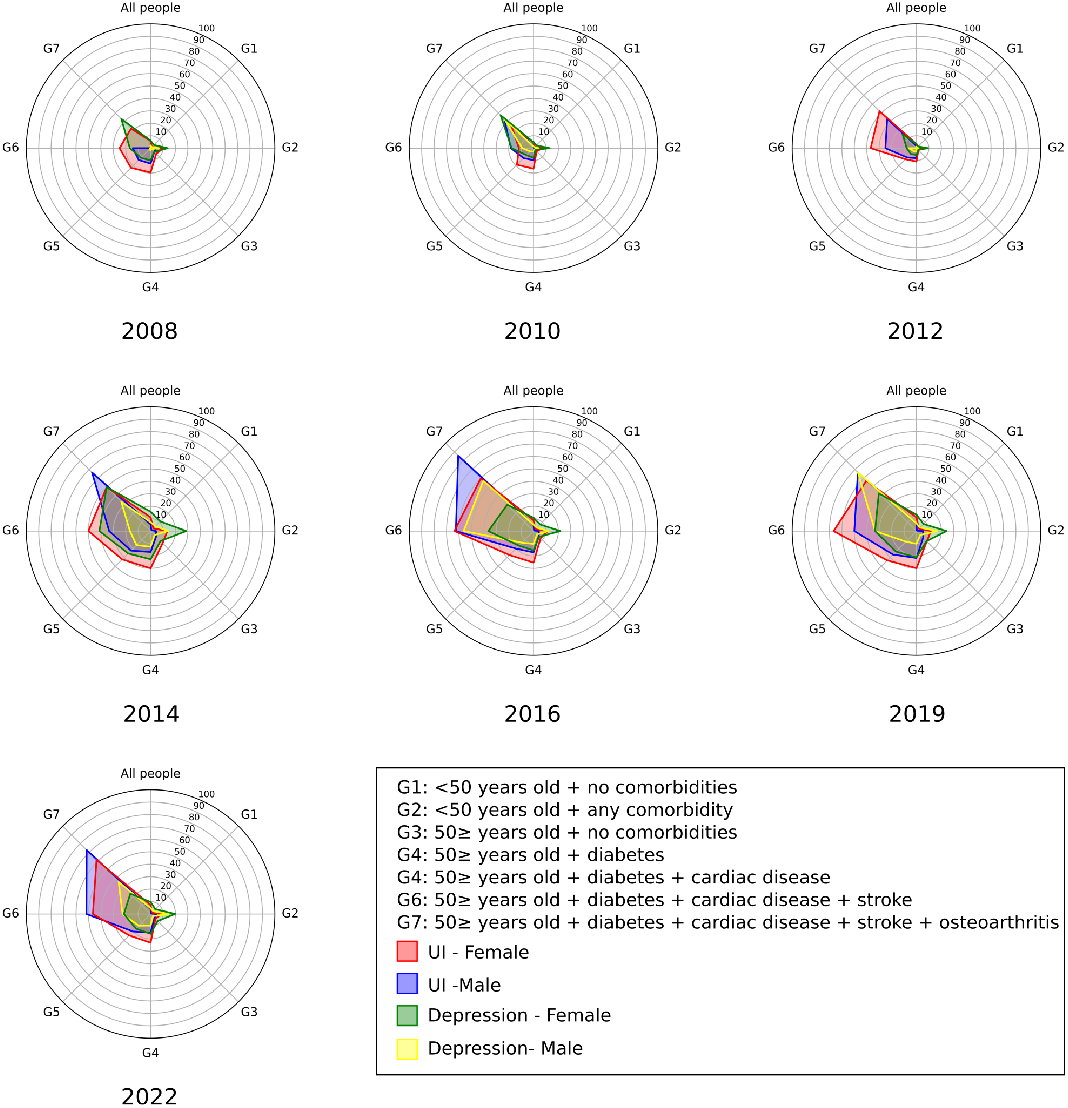
UI and depression prevalence significantly changes between years and different subgroups. Radial plots were created for each year, representing seven different subgroups categorized by age and number of comorbidities. Each radial plot corresponds to a specific year, ranging from 2008 to 2022. Within the radial plots, each circle represents a 10 percent interval.

**Figure 3.**
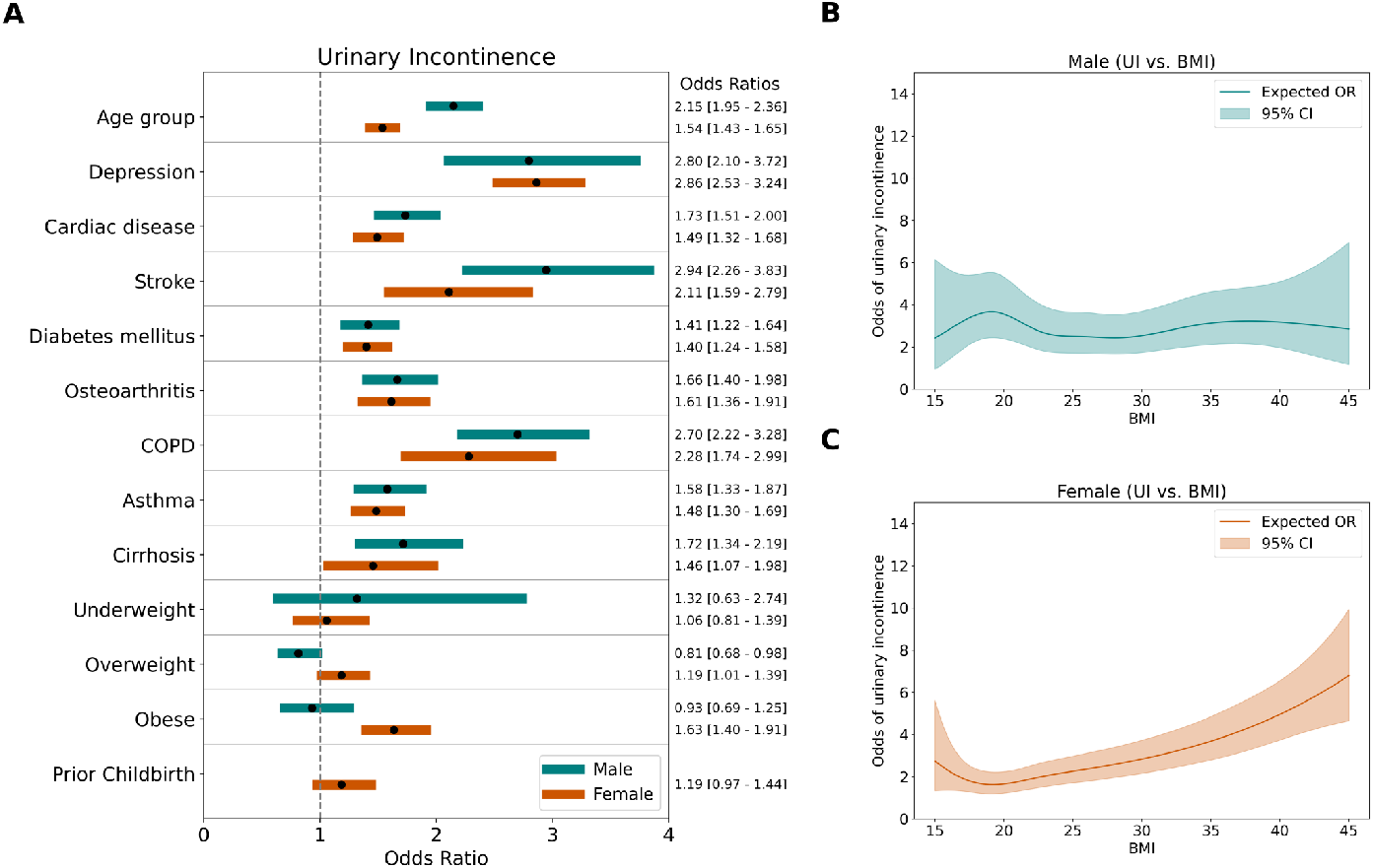
Age and chronic diseases associated with increase in urinary incontinence. Multivariable logistic regression was conducted separately for each year, with urinary incontinence as the dependent variable. Odds ratios were pooled for each independent variable. Age and all chronic conditions significantly increased the odds of urinary incontinence in both genders, while obesity was only associated with urinary incontinence in females **(A)**. Restricted cubic spline (RCS) curves were drawn to visualize the UI-BMI association in males **(B)** and females **(C)**. Solid lines represent expected odds ratio while shades show the 95% confidence interval.

Conversely, obese females had a higher likelihood of experiencing UI. Prior childbirth increases the prevalence of UI in females (OR 1.19 [0.97-1.44, 95%CI]). To thoroughly evaluate the relationship between urinary incontinence (UI) and BMI, we pooled participants from all years and conducted a restricted cubic spline (RCS) regression (Figure 3B, C). The model revealed an almost linear increasing trend for females, while a more constant trend was observed for males. However, the confidence intervals were wide at both ends due to the low number of individuals in those categories. For lifestyle and social factors, we applied both univariable and multivariable logistic regression, adjusting for age, BMI, and other comorbidities. The adjusted odds ratio for an urban lifestyle was 0.77 [0.63-0.93] for females and 0.63 [0.49-0.80] for males (Table 2). People with medium and high education levels had lower odds of having UI compared to those with low education in both genders. Marital status was not significant in females, while married males exhibited a lower adjusted odds ratio (OR 0.71 [0.59-0.84, 95We observed that as individuals’ income levels increase, median PHQ-8 scores decrease (Figure 4A), with low-income individuals exhibiting higher PHQ-8 scores. Moreover, people with low incomes experience higher rates of depression and UI, and the prevalence of both conditions decreases as income levels rise (Figure 4B). Significant differences in UI prevalence between income levels are documented in Supplementary Table 1. Additionally, the distribution of depression and UI prevalence across statistical regions (NUTS2) shows a high correlation (Figure 4E, r = 0.74, p<0.01). In addition to the strong association between UI and depression, we observed that the prevalence of both conditions follows the same trend over the years in both genders (r = 0.98, p<0.01) (Figures 4C, D and Supplementary Figure 2). Although the prevalence of UI and depression consistently decreased between 2008 and 2012, we noted a sudden surge in 2014 followed by some fluctuations. For this reason, we applied segmented regression to the interrupted time series data. We assumed an intervention occurred between 2012 and 2014 and tested its impact on the prevalence of depression and UI. The model showed good fit for both UI (Residual standard error = 0.13, Adjusted R^2^ = 0.99) and depression (Residual standard error = 0.64, Adjusted R^2^ = 0.99). Prior to 2014, there was a downward trend in the prevalence of depression and UI, indicated by the orange dashed lines. In 2014, there was a sharp increase followed by fluctuations towards a decreasing trend, although levels remained higher than before the event. Generally, the time factor correlates negatively in both the depression and UI models. Postevent, the time factor correlates negatively with depression but contributes positively to the prevalence of UI. This is also visualized by the blue lines, where depression exhibits a lower negative slope. To understand the sudden increase in the prevalence of depression and UI in 2014, we thoroughly examined nationwide events and socio-economic changes in Turkey (Figure 4F). According to data from TurkStat, Turkey’s unemployment rate began to rise in 2013, reaching 8.73%, and peaked in 2019 at 13.67%. Similarly, Turkey’s GDP per capita and GNI began to decline after 2013, indicating an economic downturn. There was a positive correlation between the unemployment rate and the prevalence of UI and depression; however, it was not statistically significant (r = 0.68, p = 0.06 for UI; r = 0.55, p = 0.129 for depression).

**Figure 4.**
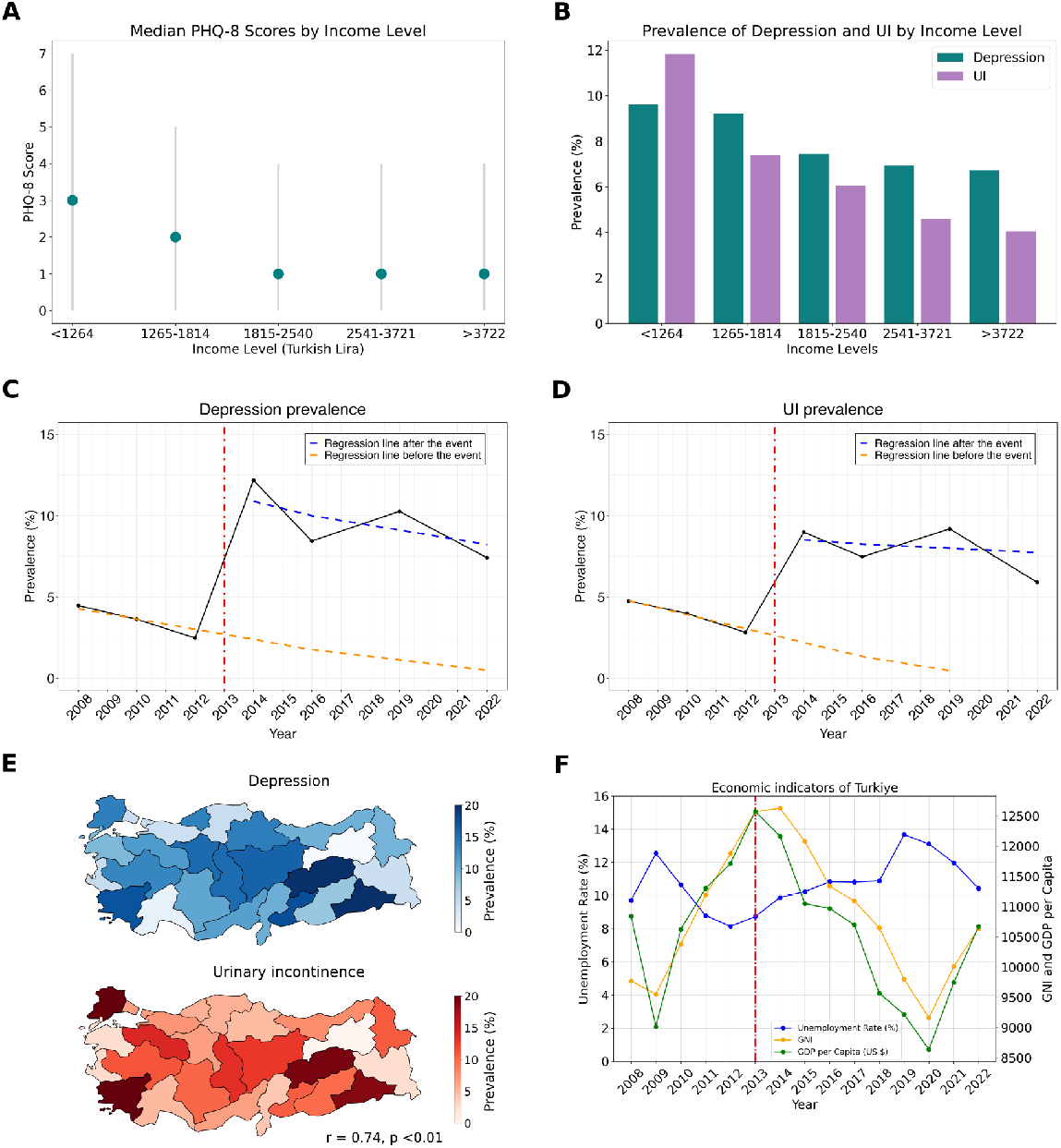
Effect of income and macroeconomic indicators on UI and depression prevalence. Median PHQ-8 scores by income level (A). Dots represent median values, while the gray line indicates the lower and upper quartiles. Prevalence of depression and urinary incontinence (UI) by income level, showing a decrease in both conditions as income increases **(B)**. Interrupted time series analysis for depression **(C)** and UI (**D)**. The vertical red line marks the event that disrupts the natural trend of the diseases. The orange dashed line depicts the underlying trend of prevalence prior to and projected after the event, assuming no disruption had occurred. The blue dashed line shows the trend of prevalence following the event. Changes in the levels between the orange and blue lines indicate the immediate impact of the event, while changes in the slopes represent its ongoing effects. Distribution of depression and UI prevalence across NUTS-2 regions **(E)**. Visualization of the unemployment rate, GNI, and GDP per capita **(F)**. Data sourced from TurkStat.

## Discussion

In the present study, we extensively analyzed the trends and associations between urinary incontinence (UI) and depression in a large, representative national sample from the Turkiye Health Interview Survey spanning from 2008 to 2022. Our findings underscore a significant and consistent association between UI and depression across different years, genders, and age groups, highlighting a robust link regardless of the underlying demographic and health status factors. Our analysis revealed that higher depression severity correlates with an increased likelihood of UI, reinforcing the potential bidirectional nature of this relationship. The age and number of comorbid conditions also emerged as critical factors, with older individuals and those with multiple chronic conditions showing higher prevalences of both UI and depression. Notably, we observed that both conditions displayed parallel trends over the study period, with a marked increase in prevalence in 2014, followed by fluctuations but remaining at elevated levels compared to the initial years of the survey. This surge aligns temporally with significant socioeconomic changes in Turkey, suggesting that broader societal stresses may influence these health outcomes.

Both cross-sectional and longitudinal studies in the current literature have consistently demonstrated a strong bidirectional association between UI and psychological discomfort, particularly depression, across both genders. Individuals with UI often experience increased levels of psychological distress, compounded by the physical limitations and social stigmatization associated with UI, while depression can further exacerbate the perception of UI severity and hinder the effective management of both UI and its comorbidities (34; 17). In line with these findings, our previous study revealed a similar pattern, where depression prevalence increased alongside UI, particularly in middle-aged women. In the present research, we provide further evidence supporting this firm relationship between UI and depression by using a large, nationally representative dataset spanning multiple years. Notably, our results not only reaffirm this well-established association but also expand on previous research by incorporating the impact of additional contributing factors such as aging, chronic conditions, and lifestyle elements. Moreover, this comprehensive analysis was done in light of macroeconomic and temporal contexts, which may reflect broader societal or economic changes influencing the trends during that 14-year period. Thus, it enhances our understanding of how these two conditions evolve together over time and across different demographic and medical subgroups. Previous studies have attributed the association between UI and chronic conditions such as diabetes and cardiovascular diseases to their role in compromising pelvic floor function (2; 35). Neurodegenerative diseases like Parkinson’s and Alzheimer’s or stroke have also been blamed for contributing to UI through both disrupted neurological pathways and the overall decline in motor control (35). Functional impairments and mobility problems are associated with UI via decrease in muscle quality (12). By conducting multivariable analysis, we present that different chronic conditions are associated with UI and increased the likelihood of having it (Figure 3).It is also observed and proven that the presence of chronic conditions, particularly when multiple comorbidities are involved, has been linked to greater psychological distress, which subsequently leads to depression (26). In parallel with the existing literature, chronic illnesses significantly impact both urinary incontinence (UI) and depression prevalence in our study population (Figure 3).

Interestingly, as shown in Figure 2, the current findings reveal that both sexes follow a similar trajectory, with no significant gender differences in the relationship between comorbidities, UI, and depression. However, a distinctive feature of this study is the critical role of age in modulating these associations. Before the age of 50, comorbidities exert a relatively minimal impact on the development of UI and depression. In contrast, after 50, the presence of multiple chronic conditions markedly elevates the risk of both UI and depression, underscoring the compounded effect of aging and comorbidities.

We observed that increase in age, having a cardiac disease, stroke and COPD had higher odds ratios for having UI in males compared to females, while obesity was significant risk factor only for females (Figure 3A). Thus, we further evaluated the gender specific relationship between UI and BMI. Previous studies have shown that obesity is a modifiable risk factor for UI. However, only limited number of studies focused on gender specific analysis for the relationship between BMI and UI. In line with a previous nationwide longitudinal study (36), we found that BMI is linearly correlated with UI in females, but not in males (Figure 3B, C). The mechanism by which obesity increases the risk of UI is explained by multiple factors (36; 37). Excess body weight leads to increased abdominal pressure, which in turn augments bladder pressure and shifts urethral position, directly contributing to stress urinary incontinence (SUI) and exacerbating symptoms of detrusor instability and overactive bladder. Furthermore, obesity induces chronic strain and mechanical stress, stretching and weakening pelvic floor muscles, nerves, and other supporting structures, thus impairing pelvic organ function. This biomechanical stress is compounded by biochemical changes; obesity is associated with systemic inflammation and oxidative stress, which promote vascular damage and sclerosis of the pelvic floor muscles and bladder tissues. These physiological changes are particularly evident in women, given their unique pelvic anatomy and factors such as pregnancy, childbirth, and menopause, which further damage pelvic tissues and diminish muscle tone and function. Increase odds of having UI in underweight men can be explained by an unmeasured covariate. Chen et. al. suggested that this covariate can be frailty, which increases disability and hospitalization and consequently UI. However, it should be acknowledged that frailty caused by underweight should have affected both genders equally. The further analysis with UI subtypes should be conducted to understand gender differences in BMI-UI relationship since the type of UI is different for males and females. Economic factors have been recognized as significant determinants of mental health for a long time. Thoits and Hannan et. al. (38), one of the first research groups, established that depression tends to inversely correlate with income, a finding supported by later empirical studies demonstrating that individuals with lower personal or household income are at an increased risk of depression (39; 40; 41). Conversely, a recent Chinese large-scale survey study revealed a U-shaped correlation where mental health issues are particularly pronounced at both low and high ends of the socioeconomic spectrum (42). In contrast, our findings, described in Figure 4A, showed that PHQ-8 scores decreased with increasing income up to a certain threshold, after which they plateaued.

Similarly, the prevalence of UI is influenced by economic conditions. Research shows that lower income and socioeconomic status contribute to higher rates of UI, partly due to limited access to healthcare resources and higher levels of psychological stress (2; 43; 44).

Not only economic but also socioeconomic factors have been shown to impact the prevalence of both UI and depression. Socioeconomic factors affecting UI consist of education, occupation, income, economic hardship and housing tenure, poverty income ratio (43; 44) and the ones affecting depression are income, marital status, economic hardship (EHQ), financial threat (FTS), financial well-being (FWBS) (42; 41; 45).

Our analysis, as depicted in 4, highlights a notable shift in the prevalence of UI and depression in Turkey between 2012 and 2014. During this period, Turkey experienced economic difficulties and social unrest. Recent studies have highlighted that socio-political events and social unrest contribute to major mental health burden, especially depression and post-traumatic stress (46; 47). These findings underscore that the observed changes in UI and depression prevalence are not solely attributable to fluctuations in GDP per capita but are likely influenced by a complex interplay of economic and sociopolitical factors. Current literature comprises several studies supporting the correlation at the individual level while indicating the intricate nature of this relationship at the country level (48; 49).

We discovered that a history of childbirth can increase the prevalence of UI among women, regardless of whether they have had a cesarean section (C-section) or a vaginal delivery. However, it was observed that vaginal delivery increases the UI prevalence more than C-section (50). Given that Turkey’s fertility rate is declining each year (Turkish Statistical Institute, 2024) and C-section rates are as high as 60% (51)—with an increasing trend—it can be expected that UI prevalence may decrease. Apart from the surge in 2014, our segmented model (Figure 4D) suggests a decrease in UI prevalence, particularly among the female population. This difference in prevalence change between genders may be explained by shifts in both fertility rates and childbirth practices.

An incidental finding in our dataset is the worrisome increase in the prevalence of Diabetes Mellitus (DM) in Turkey. DM is one of the major causes of mortality and morbidity and its prevalence is expected to rise globally (52). In our dataset, there is a linear increase in DM prevalence, rising from 6.49% to 12.06% over 14 years. Our findings support a study that analyzed all Electronic Health Records in 2020, which reported a DM prevalence of 11.12% based on lab-based diagnostic criteria (53). Nationwide, appropriate preventive, screening, and treatment strategies should be implemented.

Our results should be interpreted in the context of a few potential limitations. Being an observational study, it is inherently limited to identifying associations and correlations rather than establishing direct causation. This restricts our ability to definitively state that one factor causes another, as the relationships may be influenced by unknown or unmeasured variables. This national health survey data does not contain measures to characterize UI, or the quantity of urine lost per episode of incontinence. These are important dimensions to understand the mechanism between UI and its risk factors in both genders. The presence of chronic conditions are all patient-reported outcomes (PROs) and irrespective of a doctor’s confirmation or formal diagnosis. Additionally, while we observed a notable surge in the prevalence of depression and UI following an interruption in 2014, the observational nature of the study constrains our ability to pinpoint and fully explain the causative external factors or how they exert their influence on UI prevalence. The inability to clarify the mechanisms through which these external factors impact UI and depression underscores the need for further research.

Despite the limitations, the present study’s strengths are evident through the comprehensive and longitudinal analysis of data collected over seven distinct years, involving a large and diverse sample size that reflects the general population of Turkiye. Such a longitudinal approach allowed us to not only confirm the high correlation between UI and depression but also to explore the interaction with various other chronic conditions and demographic factors. The extensive list of variables considered in our analysis—ranging from biological factors like age, body mass index (BMI), and a spectrum of chronic conditions, to socioeconomic factors including education levels, marital status, and urban vs. rural living—enhanced our understanding of the complex interplay between these elements and their impact on UI. This broad variable spectrum strengthens the study’s ability to provide a holistic view of the health landscape over an extended period, making it a valuable resource for public health assessments and policymaking.

## Conclusion

Conducting nationwide health surveys in regular time intervals is an effective strategy to capture much needed data of disease prevalence in a spatiotemporal manner. They allow us to guide in describing the current state of diseases and in assessing the impact of our health policies and interventions on various levels. The present study is a good example of such utilization of national survey data, showing the need for a more holistic approach towards the diagnosis, treatment, and follow-up of urinary incontinence as our study suggests a complex association of UI with depression and patient characteristics, which should be taken into account in disease prevention and management.

## Funding

MK received partial financial support from TUBITAK Directorate of Science Fellowships and Grant Programmes (BIDEB)-2232 International Fellowship for Outstanding Researchers (Award No: 118C306).

## Data Availability

The data used in this study were obtained from the Turkish Statistical Institute (TurkStat), but the accessibility of these data is limited due to certain constraints. The data were licensed specifically for this study and are not publicly available. However, authors can provide the data upon a reasonable request and with included permission from TurkStat, and the Regional Ethical Committee.

## Acknowledgment

We thank the Turkish Statistical Institute (TurkStat) for data sharing

## Notes

### Competing Interest Statement

The authors have declared no competing interest.

### Author Declarations

Ethical approval was granted by the Istanbul Medipol University Ethics Committee (Application number: 10840098-604.01.01-E.53819). All procedures were carried out in compliance with the Declaration of Helsinki.

