## Supplementary_file for "Exploring the Association Between Urinary Incontinence and Depression Based on a Series of Large-Scale National Health Studies in Turkiye"

Muhammed Furkan Dasdelen[1], Zehra Betul Dasdelen[1], Furkan Almas[1], Beyza Cokkececi[3], Pilar Laguna[1,2], Jean de la Rosette[1,2], Mehmet Kocak[1,4*]
[1] International School of Medicine, Istanbul Medipol University, Istanbul, Turkiye
[2] Department of Urology, Istanbul Medipol University, Istanbul, Turkiye
[3] Faculty of Engineering and Natural Sciences, Sabanci University, Istanbul, Turkiye
[4] Department of Biostatistics and Medical Informatics, Istanbul Medipol University, Istanbul, Turkiye

*Correspondence: Muhammed Furkan DASDELEN

Address: Kavacik Mah. Ekinciler Cad. No:19, 34810, Beykoz/Istanbul

### Supplementary File

### Supplementary Figures

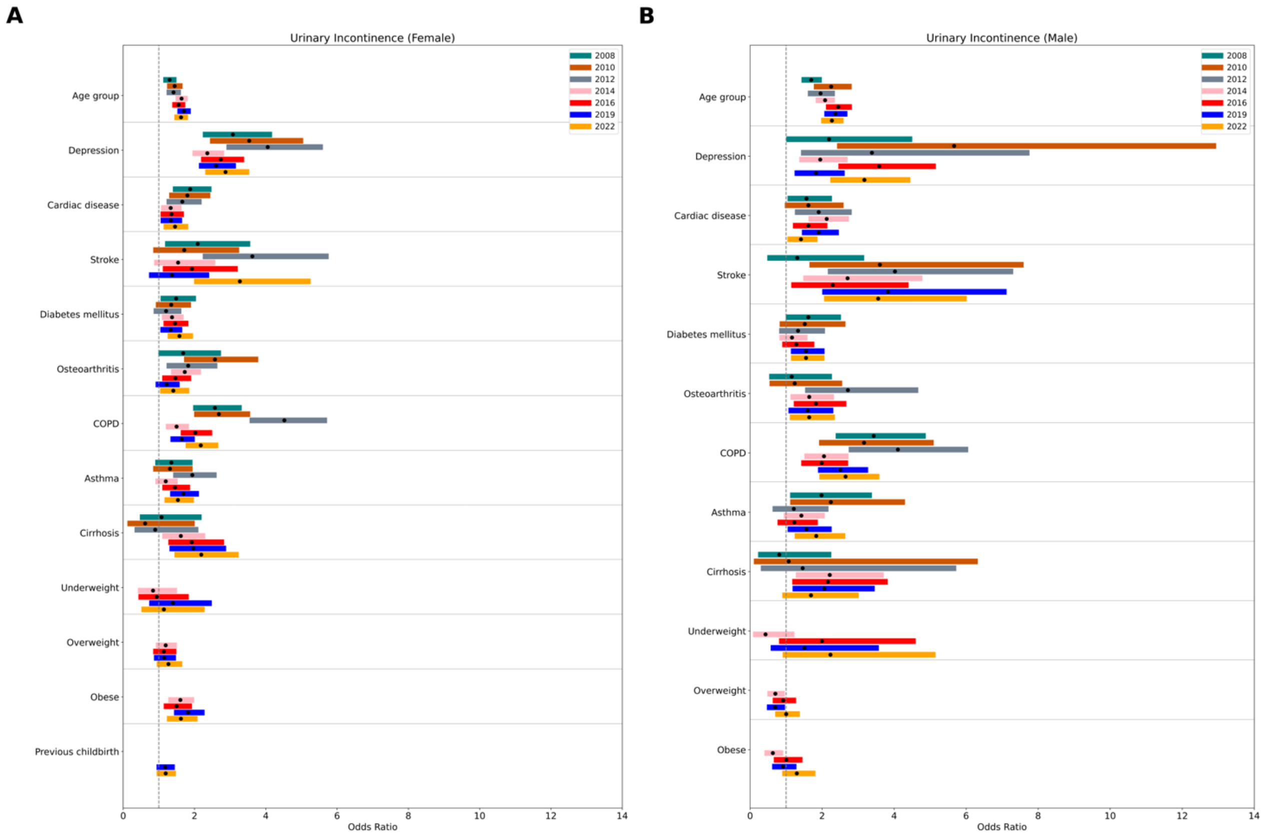

**Supplementary Figure 1: Detailed UI odds ratios by year.** Separate multivariable logistic regression models were fitted for each year across both sexes. Different colors represent different years, as specified in the legend. Black dots indicate the odds ratios, and colored bars show the 95% confidence intervals.

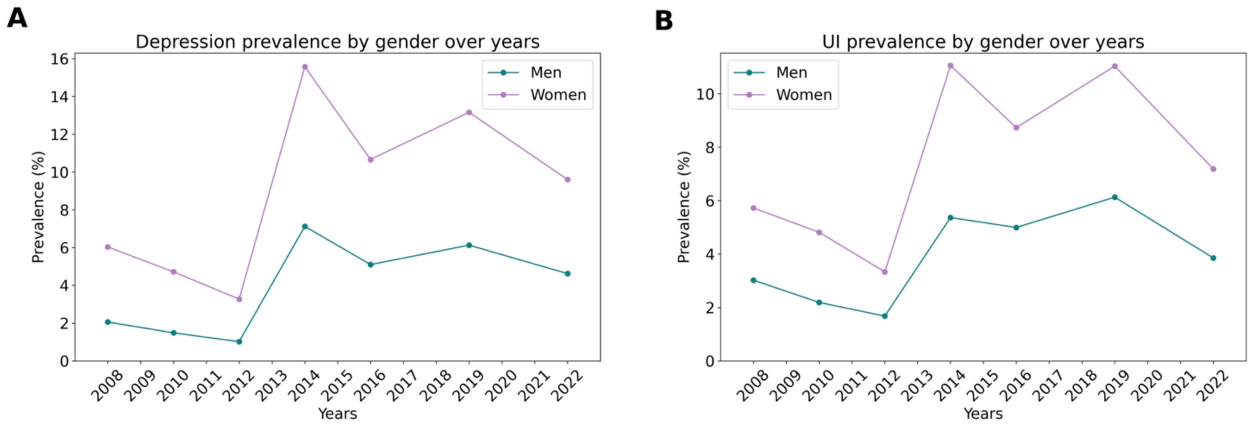

**Supplementary Figure 2. UI and depression prevalence follows the similar trend in both sexes over years.** Depression **(A)** and UI prevalence **(B)** were depicted separately for both genders. Females have higher rate of both depression and UI than males. The trend of prevalence change in depression and UI follows similar pattern in both genders.

### Supplementary Tables

**Supplementary Table 1:** UI prevalence in different income levels

| Income Level | 2008<br>N (%) | 2010<br>N (%) | 2012<br>N (%) | 2014<br>N (%) | 2016<br>N (%) | 2019<br>N (%) | 2022<br>N (%) |
|---|---|---|---|---|---|---|---|
| **1** | 71 (7.6%) | 55 (5.8%) | 32 (5.9%) | 754 (12.8%) | 434 (11.8%) | 43 (18.6%) | 18 (12.0%) |
| **2** | 68 (8.8%) | 60 (6.0%) | 15 (2.9%) | 303 (7.9%) | 347 (7.4%) | 21 (14.1%) | 10 (11.1%) |
| **3** | 25 (4.1%) | 46 (4.0%) | 12 (3.0%) | 225 (7.2%) | 191 (6.1%) | 22 (17.9%) | 3 (4.9%) |
| **4** | 25 (5.5%) | 71 (5.2%) | 16 (2.5%) | 206 (6.3%) | 135 (4.6%) | 20 (23.3%) | 9 (19.6%) |
| **5** | 29 (6.6%) | 38 (3.2%) | 30 (3.0%) | 131 (4.4%) | 112 (4.0%) | 15 (24.2%) | 21 (18.1%) |
| **6** | 20 (4.2%) | 56 (4.2%) | 12 (1.2%) | | | 10 (5.7%) | 4 (8.2%) |
| **7** | 8 (2.2%) | 17 (2.2%) | 20 (2.3%) | | | 18 (10.8%) | 14 (15.2%) |
| **8** | 21 (4.9%) | 45 (2.7%) | 27 (2.0%) | | | 8 (9.2%) | 17 (13.2%) |
| **9** | 5 (2.6%) | 24 (2.3%) | 22 (2.1%) | | | 8 (7.0%) | 8 (7.3%) |
| **10** | 12 (4.3% | 33 (2.4%) | 26 (1.9%) | | | 5 (6.6%) | 4 (3.9%) |
| **11** | | | | | | 10 (7.0%) | 14 (7.5%) |
| **12** | | | | | | 4 (6.8%) | 13 (4.8%) |
| **13** | | | | | | 5 (6.8%) | 11 (5.8%) |
| **14** | | | | | | 12 (10.6%) | 1 (0.8%) |
| **15** | | | | | | 14 (13.7%) | 13 (8.7%) |
| **16** | | | | | | 8 (7.1%) | 13 (7.0%) |
| **17** | | | | | | 7 (9.9%) | 6 (3.4%) |
| **18** | | | | | | 6 (8.1%) | 15 (4.2%) |
| **19** | | | | | | 3 (3.5%) | 7 (2.7%) |
| **20** | | | | | | 3 (3.0%) | 13 (2.3%) |
| **X** | 38.35 | 58.59 | 38.75 | 234.67 | 197.92 | 73.50 | 112.59 |
| **P value*** | <0.001 | <0.001 | <0.001 | <0.001 | <0.001 | <0.001 | <0.001 |

* After Bonferroni correction, P value less than 0.007 was considered significant

**Supplementary Table 2.** Classification of income levels by years

| Income Level | **2008 N (%)** | **2010 N (%)** | **2012 N (%)** | **2014 N (%)** | **2016 N (%)** | **2019 N (%)** | **2022 N (%)** |
|---|---|---|---|---|---|---|---|
| **1** | <350 | <350 | <350 | <1080 | <1264 | <992 | <1646 |
| **2** | 351-500 | 351-500 | 351-500 | 1081-1550 | 1265-1814 | 993 - 1298 | 1647 - 2212 |
| **3** | 501-620 | 501-620 | 501620 | 1551-2710 | 1815-2540 | 1299 - 1500 | 2213 - 2570 |
| **4** | 621-750 | 621-750 | 621-750 | 2711-3180 | 2541-3721 | 1501 - 1668 | 2571 - 2883 |
| **5** | 751-900 | 751-900 | 751-900 | >3181 | >3722 | 1669 - 1849 | 2884 - 3172 |
| **6** | 901-1100 | 901-1100 | 901-1100 | | | 1850 - 2025 | 3173 - 3472 |
| **7** | 1101-1300 | 1101-1300 | 1101-1300 | | | 2026 - 2214 | 3473 - 3774 |
| **8** | 1301-1700 | 1301-1700 | 1301-1700 | | | 2215 - 2424 | 3775 - 4111 |
| **9** | 1701-2300 | 1701-2300 | 1701-2300 | | | 2425 - 2656 | 4112 - 4462 |
| **10** | >2301 | >2301 | >2301 | | | 2657 - 2892 | 4463 - 4852 |
| **11** | | | | | | 2893 - 3144 | 4853 - 5268 |
| **12** | | | | | | 3145 - 3398 | 5269 - 5713 |
| **13** | | | | | | 3399 - 3695 | 5714 - 6199 |
| **14** | | | | | | 3696 - 4052 | 6200 - 6794 |
| **15** | | | | | | 4053 - 4492 | 6795 - 7510 |
| **16** | | | | | | 4493 - 5052 | 7511 - 8372 |
| **17** | | | | | | 5053 - 5784 | 8373 - 9539 |
| **18** | | | | | | 5785 - 6890 | 9540 - 11356 |
| **19** | | | | | | 6891 - 8912 | 11357 - 14812 |
| **20** | | | | | | >8913 | >14813 |

### Supplementary Codes

#### Code for multivariable logistic regression:

```
import pandas as pd
import numpy as np
import statsmodels.api as sm
from sklearn.preprocessing import LabelEncoder
import statsmodels.formula.api as smf
import os

def save_model(year,df,indep_var,dep_var_string,gender,save_root):
  formula = indep_var + " ~ " + dep_var_string
  model = smf.logit(formula = formula,data=df).fit()
  print(model.summary())
  odds_ratios = pd.DataFrame(
    {
      "OR": model.params,
      "Lower CI": model.conf_int()[0],
      "Upper CI": model.conf_int()[1],
    }
  )
  odds_ratios = np.exp(odds_ratios)
  odds_ratios["p_values"] = model.pvalues

  odds_ratios.reset_index().to_excel(os.path.join(save_root,f'odds_ratio_{year}_{gender}.xlsx'),index =
False)
  odds_ratios

years = ['2008','2010','2012','2014','2016','2019','2022']

root = '/Users/furkan/Desktop/Data/Engineering/Biostatistics/Turkish Health Studies/Turkish Health
Studies/'

for year in years:
  csv_path = root + year+'_cleaned_data.csv'
  df = pd.read_csv(csv_path)

  #df['bmi_cat'].replace('normal',0,inplace=True)
  #df['bmi_cat'].replace('underweight',1,inplace=True)
  #df['bmi_cat'].replace('overweight',2,inplace=True)
  #df['bmi_cat'].replace('obese',3,inplace=True)
  #df['bmi_cat'].replace('na',4,inplace=True)

  df['YAS_GRUBU'].replace('15-24',0,inplace=True)
  df['YAS_GRUBU'].replace('25-34',1,inplace=True)
  df['YAS_GRUBU'].replace('35-44',2,inplace=True)
```

```
    df['YAS_GRUBU'].replace('45-54',3,inplace=True)
    df['YAS_GRUBU'].replace('55-64',4,inplace=True)
    df['YAS_GRUBU'].replace('65-74',5,inplace=True)
    df['YAS_GRUBU'].replace('75+',6,inplace=True)
    df['YAS_GRUBU'].replace(6,5,inplace=True)

    df_men = df[df.cinsiyet==0]
    df_women = df[df.cinsiyet==1]

    save_root = 'odds_ratios_biologic'
    print('Year: ', year)
    indep_var = "ui"
    # for including bmi
    if year in ['2014','2016','2019','2022']: # to do add 2022
        dep_var_string = "YAS_GRUBU + depression + asthma + copd + cardiac_disease + stroke +
osteoarthritis +  diabetes + cirrhosis + C(bmi_cat)"
    else:
        dep_var_string = "YAS_GRUBU + depression + asthma + copd + cardiac_disease + stroke +
osteoarthritis +  diabetes + cirrhosis"
    save_model(year,df_men,indep_var,dep_var_string,'men',save_root)
    save_model(year,df,indep_var,dep_var_string + ' + cinsiyet','all',save_root)
    if year in ['2019','2022']: # to do add 2022
        dep_var_string = dep_var_string + ' + pregnancy'
    save_model(year,df_women,indep_var,dep_var_string,'women',save_root)
```

**Code for pooling odds ratios:**

```
library(metafor)
library(readxl)
library(writexl)

read_and_rename <- function(file_path) {
  data <- read_excel(file_path)
  colnames(data) <- c("index", "OR", "Lower_CI", "Upper_CI", "p_values")
  return(data)
}

path <- "odds_ratios_biologic/"
# Include all years
years <- c(2008,2010,2012,2014,2016,2019,2022)

# Change men or women
pattern <- paste0("odds_ratio_", years, "_men.*\\.xlsx$")  # Creating the pattern

# Combining patterns with OR operator to match any of the specified years
full_pattern <- paste(pattern, collapse="|")

# List all files matching the pattern
file_names <- list.files(path, pattern = full_pattern, full.names = TRUE)

# Read the data from Excel files
data_list <- lapply(file_names, read_and_rename)

prepare_data <- function(df) {
  df$logOR <- log(df$OR)
  df$varLogOR <- (log(df$Upper_CI) - log(df$Lower_CI)) / (2 * 1.96)^2  # Approximation of variance
  return(df)
}
data_list <- lapply(data_list, prepare_data)

# Combine all data into one dataframe
combined_data <- do.call(rbind, data_list)

# Perform the meta-analysis for each variable
unique_vars <- unique(combined_data$index)

results <- lapply(unique_vars, function(var) {
  dat <- combined_data[combined_data$index == var, ]
  rma_res <- rma(yi = logOR, sei = sqrt(varLogOR), data = dat, method = "REML")
    temp_df <- data.frame(
    Variable = var,
    OR = exp(rma_res$b),
```

```
    Lower_CI = exp(rma_res$b - 1.96 * rma_res$se),
    Upper_CI = exp(rma_res$b + 1.96 * rma_res$se),
    p_value = rma_res$pval
  )
  return(temp_df)
})
results_df <- do.call(rbind, results)

# Save the results to an Excel file, change men or women
write_xlsx(results_df, paste0(path, "pooled_ORs_men.xlsx"))
```

#### Code for segmented regression:

```
library(tidyverse)
library(scales)
library(prais)
library(DT)
data <- read_csv(r'(aligned_data.csv)')

data <- data %>%
  # Define variables time, event (intervention), and time_event (time after intervention)
  mutate(time = row_number())%>%
  mutate(event = if_else(time >=4,1,0))%>%
  mutate(time_event = case_when(event == 1 ~ row_number()-3, TRUE~0))

ggplot(data = data)+
  # Plotting line chart
  geom_point(aes(x = Year, y = depression_prev),color ='orange')+
  geom_line(aes(x = Year, y = depression_prev),group =1,color ='orange')+
  geom_point(aes(x = Year, y = ui_prev),color ='blue')+
  geom_line(aes(x = Year, y = ui_prev),group =1,color ='blue')+
  # Plotting vertical line to segmented the time series (before and after event)
  geom_vline(xintercept = 2014, linetype="dotdash", color ='red',size=1.2)+
  # Following code is aesthetic adjustment
  scale_y_continuous(limits = c(0,0.25),labels = comma)+
  xlab('Time')+
  ylab('Prevalence')+
  labs(title = 'Time Series Depression UI')+
  theme_bw()+
  theme(axis.text.x = element_text(angle = 90, vjust = 0.3,size = 16),
      axis.text = element_text(size=16),
      axis.title = element_text(size = 16),
      plot.title = element_text(size=20))

# Fit regression for depression
model <- prais::prais_winsten(depression_prev~ time + event + time_event,
                    index ='time',
                    data = data)
# Fit regression for UI
model2 <- prais::prais_winsten(ui_prev~ time + event + time_event,
                    index ='time',
                    data = data)

# Add model coefficients to tha table
data<- data%>%
  mutate(factual_trend= model$coefficients[1]+
       model$coefficients[2]*time+
```

```
        model$coefficients[3]*event+
        model$coefficients[4]*time_event
  )%>%
  mutate(counter_fact = model$coefficients[1]+
        model$coefficients[2]*time)

# Add model coefficients to tha table
data<- data%>%
  mutate(factual_trend2= model2$coefficients[1]+
        model2$coefficients[2]*time+
        model2$coefficients[3]*event+
        model2$coefficients[4]*time_event
  )%>%
  mutate(counter_fact2 = model2$coefficients[1]+
        model2$coefficients[2]*time)

# plot regression model for Depression
plot1 <- ggplot(data = data)+
  ##Plotting line chart
  geom_point(aes(x = Year, y = depression_prev),size=2)+
  geom_line(aes(x = Year, y = depression_prev),group =1,size=0.8)+
  ##Plotting vertical line to segmented the time series (before and after COVID-19)
  geom_vline(xintercept = 2013, linetype="dotdash", color ='red',size=1.2)+
  ##plotting factual trend
  geom_line(data = data%>%filter(time>=4)
         ,aes(x = Year, y = factual_trend,color = 'Regression line after the event'),group =1, linetype="dashed",
size=1.2)+
  ##plotting counter-factual trend
  geom_line(aes(x = Year, y = counter_fact,color = 'Regression line before the event'),group =1,
linetype="dashed", size=1.2)+
  ## Setting x axis labels
  scale_x_continuous(breaks = seq(2008, 2022, by = 1)) +
  ##Following code is aesthetic adjustment
  scale_y_continuous(limits = c(0,15),labels = comma)+
  scale_color_manual(values = c('blue', 'darkorange'))+
  xlab('Year')+
  ylab('Prevalence (%)')+
  labs(title = 'Depression prevalence',color='')+
  theme_bw()+
  theme(axis.text.x = element_text(angle = 45, vjust = 0.6,size = 20,color='black'),
      axis.text.y = element_text(size = 20,color='black'),
      axis.title.x = element_text(size = 20, vjust = 0.2,color = "black"),
      axis.title.y = element_text(size = 20, color = "black"),
      axis.text = element_text(size=20),
      axis.title = element_text(size = 20),
      plot.title = element_text(size=24,hjust = 0.5),
      panel.border = element_rect(colour = "black", fill = NA, linewidth = 0.8),
      legend.position = c(0.75,0.9),
```

```
      legend.text = element_text(size=16),
      legend.background = element_rect(fill = "white", colour = "black",linewidth = 0.4),
      legend.title = element_blank())

ggsave("plot_depression.png", plot = plot1, width = 10, height = 7, dpi = 300)

# plot regression model for UI
plot2 <- ggplot(data = data)+
  ##Plotting line chart
  geom_point(aes(x = Year, y = ui_prev),size=2)+
  geom_line(aes(x = Year, y = ui_prev),group =1,size=0.8)+
  ##Plotting vertical line to segmented the time series (before and after COVID-19)
  geom_vline(xintercept = 2013, linetype="dotdash", color ='red',size=1.2)+
  ##plotting factual trend
  geom_line(data = data%>%filter(time>=4)
        ,aes(x = Year, y = factual_trend2,color = 'Regression line after the event'),group =1,
linetype="dashed", size=1.2)+
  ##plotting counter-factual trend
  geom_line(aes(x = Year, y = counter_fact2,color = 'Regression line before the event'),group =1,
linetype="dashed", size=1.2)+
  ## Setting x axis labels
  scale_x_continuous(breaks = seq(2008, 2022, by = 1)) +
  ##Following code is aesthetic adjustment
  scale_y_continuous(limits = c(0,15),labels = comma)+
  scale_color_manual(values = c('blue', 'darkorange'))+
  xlab('Year')+
  ylab('Prevalence (%)')+
  labs(title = 'UI prevalence',color='')+
  theme_bw()+
  theme(axis.text.x = element_text(angle = 45, vjust = 0.6,size = 20,color='black'),
      axis.text.y = element_text(size = 20,color='black'),
      axis.title.x = element_text(size = 20, vjust = 0.2,color = "black"),
      axis.title.y = element_text(size = 20, color = "black"),
      axis.text = element_text(size=20),
      axis.title = element_text(size = 20),
      plot.title = element_text(size=24,hjust = 0.5),
      panel.border = element_rect(colour = "black", fill = NA, linewidth = 0.8),
      legend.position = c(0.75,0.9),
      legend.text = element_text(size=16),
      legend.background = element_rect(fill = "white", colour = "black",linewidth = 0.4),
      legend.title = element_blank())

ggsave("plot_UI.png", plot = plot2, width = 10, height = 7, dpi = 300)
```

#### Code for figure 3:

```
import matplotlib.pyplot as plt
import numpy as np
import pandas as pd
import os

def reorder_df(df):
    new_order = ['YAS_GRUBU', 'depression', 'cardiac_disease', 'stroke', 'diabetes', 'copd', 'osteoarthritis',
            'asthma', 'cirrhosis','C(bmi_cat)[T.1]','C(bmi_cat)[T.2]','C(bmi_cat)[T.3]','pregnancy']
    new_names = ['Age group', 'Depression', 'Cardiac disease', 'Stroke', 'Diabetes mellitus',
              'Osteoarthritis', 'COPD','Asthma','Cirrhosis','Underweight','Overweight','Obese','Prior Childbirth']

    name_map = dict(zip(new_order, new_names))
    # Apply new names
    df['Variable'] = df['Variable'].map(name_map)

    # Set the 'index' column as the DataFrame index
    # Reindex the DataFrame
    df.set_index('Variable', inplace=True)
    df = df.reindex(new_names)
    # Reset the index if you want 'index' back as a column
    df.reset_index(inplace=True)
    return df

colors = ['#008080', '#cc5500']
genders = ['men','women']
map_name = {'men':'Male',
        'women':'Female'}
colors_dict = dict(zip(genders, colors))

# Read data
bar_interval = 8
line_interval = 20
count = 16
y_values_confidence = {}
data_frames = {}
root_odds = 'odds_ratios_biologic'
for gend in genders:
    csv_path = os.path.join(root_odds, f'pooled_ORs_{gend}.xlsx')
    data_frames[gend] =
reorder_df(pd.read_excel(csv_path).drop([0]).reset_index().drop(columns=['index']))

for gend in genders:
    y_values_confidence[gend] = np.arange(len(data_frames[genders[0]])) * line_interval + count
    count += bar_interval

# Setting up the plot
```

```
plt.figure(figsize=(8, 10))
plt.rcParams.update({'font.size': 16})

# Set tick intervals
y_ticks = np.arange(len(data_frames[genders[0]])) * line_interval +bar_interval*2.5

for i in range(len(data_frames[genders[0]])):
   for gend in genders:
      df_forest = data_frames[gend]
      # Plot confidence intervals
      plt.plot([df_forest.iloc[i]['Lower_CI'], df_forest.iloc[i]['Upper_CI']],
             [y_values_confidence[gend][i], y_values_confidence[gend][i]], color=colors_dict[gend],
linewidth=8, label=map_name[f'{gend}'] if i == 0 else "")
      # Plot OR value
      plt.scatter(df_forest.iloc[i]['OR'], y_values_confidence[gend][i], color='black', zorder=5)

# Adding horizontal lines for separation
for i in np.arange(min(y_ticks)+(line_interval/2), max(y_ticks), line_interval):
   plt.axhline(y=i - 0.8, color='grey', linestyle='-', linewidth=0.5, xmin=0.00, xmax=1)

plt.xlim(0, 4)
plt.xticks(np.arange(0, 5, 1))
plt.yticks(y_ticks, data_frames[genders[0]]['Variable'])
plt.axvline(x=1, color='grey', linestyle='--')
plt.xlabel('Odds Ratio')
plt.title("Urinary Incontinence")
plt.gca().invert_yaxis()  # Inverting y-axis for better readability
plt.legend(loc='lower right', fontsize=14)
plt.tight_layout()
plt.savefig('forest_plot_pooled.png', dpi=600)
plt.show()
```

**Code for supplementary figure 1:**

```
import matplotlib.pyplot as plt
import numpy as np
import pandas as pd
import os

years = ['2008','2010','2012','2014','2016','2019','2022']
colors = ['#008080', '#cc5500', '#708090', '#ffb6c1', 'red', 'blue','orange']

colors_dict = dict(zip(years, colors))

# Read data
bar_interval = 8
line_interval = 70
count = 0
y_values_confidence = {}
data_frames = {}
root_odds = 'odds_ratios_biologic'
for year in years:
    csv_path = os.path.join(root_odds, f'odds_ratio_{year}_women.xlsx')
    data_frames[year] =
reorder_df_gender(pd.read_excel(csv_path).drop([0]).reset_index().drop(columns=['level_0']))

for year in years:
    y_values_confidence[year] = np.arange(len(data_frames[years[0]])) * line_interval + count
    count += bar_interval

# Setting up the plot
plt.figure(figsize=(15, 20))
plt.rcParams.update({'font.size': 16})

# Set tick intervals
y_ticks = np.arange(len(data_frames[years[0]])) * line_interval +bar_interval*2.5

for i in range(len(data_frames[years[0]])):
    for year in years:
        df_forest = data_frames[year]
        # Plot confidence intervals
        plt.plot([df_forest.iloc[i]['Lower CI'], df_forest.iloc[i]['Upper CI']],
                 [y_values_confidence[year][i], y_values_confidence[year][i]], color=colors_dict[year],
linewidth=10, label=f'{year}' if i == 0 else "")
        # Plot OR value
        plt.scatter(df_forest.iloc[i]['OR'], y_values_confidence[year][i], color='black', zorder=5)

# Adding horizontal lines for separation
for i in np.arange(min(y_ticks)+(line_interval/2), max(y_ticks), line_interval):
    plt.axhline(y=i - 0.8, color='grey', linestyle='-', linewidth=0.5, xmin=0.00, xmax=1)
```

```
plt.xlim(0, 14)
plt.xticks(np.arange(0, 15, 2))
plt.yticks(y_ticks, data_frames[years[0]]['index'])
plt.axvline(x=1, color='grey', linestyle='--')
plt.xlabel('Odds Ratio')
plt.title("Urinary Incontinence (Female)")
plt.gca().invert_yaxis()  # Inverting y-axis for better readability
plt.legend(loc='upper right', fontsize=14)
plt.tight_layout()
plt.savefig('forest_plot_women.png', dpi=600)
plt.show()
```
